# Disaster Education and Training Programs for Enhancing Community Preparedness: A Systematic Review from the Nursing Perspective

**DOI:** 10.1101/2025.09.16.25335950

**Authors:** Widya Addiarto, Moses Glorino Rumambo Pandin, Ah Yusuf, Rizka Yunita

## Abstract

**Background:** The high frequency of disaster occurrences in various regions has had a significant impact on affected communities, including in Indonesia. Each year, the disasters exacerbate the conditions of the population, causing discomfort and posing threats to physical and mental health, as well as increasing material losses. In light of this situation, it is crucial for nurses, through their philosophical perspective, to enhance community preparedness behaviour through disaster education and training program that is accessible to all components of society, thereby minimizing the risks and impacts of disasters.

**Objective:** This literature review aimed to examine the effectiveness disaster education and training, towards individual and community preparedness behaviour drawing on studies published between 2020 and 2025.

**Method:** Guided by the PRISMA framework, a systematic search was conducted in Scopus, ProQuest, EBSCO Host, and Web of Science. Eligible studies included disaster education and training program, individual or community responders and outcome preparedness behaviour on disaster. Thirteen studies met the inclusion criteria, encompassing quasi experimental, cross-sectional, pilot study, and randomized controlled trial designs

**Results:** Across the reviewed studies, disaster education and training are critical components of effective disaster risk reduction strategies. The studies reviewed highlight the positive impact of culturally sensitive, community-based training models in enhancing disaster preparedness. Moreover, the involvement of local leaders, schools, and healthcare professionals-particularly nurses-emerges as a key factor in increasing disaster resilience.

**Conclusion:** The implementation of disaster education and training programs has demonstrated a significant improvement in preparedness behaviour within disaster-prone areas. These findings highlight the critical role of integrating disaster education and into community initiatives, as the program effectively empowers individuals to respond more efficiently to emergencies. The involvement of nurses in these programs is crucial, as they play a key role in delivering education and training within communities. Their expertise in healthcare and community engagement makes them essential in enhancing disaster preparedness and fostering resilience.

## INTRODUCTION

Natural disasters are events that frequently occur in various regions around the world, including Indonesia (Dewi et al., 2023). Natural disasters not only cause material losses but also damage infrastructure, threaten human safety, and have profound psychological impacts on affected communities (Rahman et al., 2024). In Indonesia, many areas are prone to flooding, such as coastal regions and lowlands, which have high vulnerability levels due to extreme rainfall, climate change, and environmental degradation. Several efforts have been made by the government in collaboration with the community through the provision of gathering points during flood events. However, these initiatives have not been effectively implemented across all residents due to the limited dissemination of information, resulting in the potential for recurring disaster threats. In fact, a significant portion of the community still lacks the necessary understanding or skills to address disaster risks, particularly in disaster-prone areas (Gumelar et al., 2020). Consistent with this perspective, Addiarto dan Kusyairi (2025) argue that the majority of communities in disaster-prone areas still exhibit low disaster preparedness levels, indicating that they are not yet aware of how to prepare for and take preventive measures against floods, nor do they know how to execute swift and safe evacuations when rainfall reaches high levels. Therefore, it is essential to involve nurses in disaster preparedness within the community, including providing education and training to individuals and the surrounding community in disaster-prone areas to enhance individual and community preparedness. This is necessary because one of the primary areas of nursing practice is within the community.

Disaster preparedness has become critically important in minimizing the negative impacts of such events (Nababan et al., 2024). According to Bakic dan Ajdukovic, (2021) preparedness attitudes and awareness are essential for all levels of society, considering the substantial losses in terms of human casualties and material damages that occur during disasters. It is expected that preparedness is not only emphasized for adults, but also that children must maintain a high level of awareness regarding disaster events. Therefore, the involvement of all sectors, including government, academia, and BPBD organizations, is necessary to implement preventive measures in disaster management. This can be achieved through comprehensive disaster education, which can improve preparedness behaviour and awareness, thereby reducing disaster risks (Dewabrata et al., 2023). Enhancing preparedness should not only focus on technical aspects, but also encompass biological, psychological, social, cultural, and spiritual dimensions, all of which contribute to community resilience in the face of disaster. One effective strategy is to provide comprehensive disaster education from an early age within the family context (Al Thobaity, 2024).

Individuals and families are the smallest units within society and play a central role in disaster mitigation and response (Guo et al., 2021). Through comprehensive and structured education, knowledge, skills, and preparedness behaviour can be developed (Kurata et al., 2023). In line with this perspective, Sofyana et al., (2024) dan Noor et al., (2023) argue that comprehensive disaster education programs from a nursing viewpoint are expected to serve as an effective strategy for fostering more resilient individuals, families, and communities. This approach aims to reduce the impact of disasters and accelerate recovery in the future (Sofyana et al., 2024; Noor et al., 2023). Therefore, based on the outlined issues, this article aims to directly identify how comprehensive disaster education and training program can enhance individual and community preparedness from the perspective of nursing philosophy, through evidence-based practice. This literature review aims to examine the effectiveness of disaster education and training program towards community preparedness behaviour, focusing on studies published between 2020 and 2025.

## METHOD

### Study Design

The process of literature screening, including, and reporting was based on the guidelines of PRISMA. The search was conducted in Scopus, ProQuest, EBSCO Host and Web of Science (WoS) databases from 2020 to 2025.

### Inclusion and Exclusion Criteria

Eligibility criteria in this study included inclusion and exclusion criteria. The inclusion criteria were as follows: Programs or interventions that focus on disaster education and training, including but not limited to simulation exercises, tabletop drills, community-based participatory approaches, and theoretical-based curricula, research involving nurses, nursing students, or healthcare professionals engaged in disaster education and training programs. The exclusion criteria were as follows: Programs not specifically designed for disaster education or training, or those not led or facilitated by nursing professionals and research focusing on non-healthcare professionals.

### Searching Strategy

The literature search used Scopus, PubMed, Web of Science (WoS), Science Direct, and Ebsco databases from 2020 to 2025 using English. The keywords in the search were (disaster OR disaster preparedness) AND (disaster education OR disaster training program) AND (nursing perspective) AND (preparedness). The identification, screening, and inclusion procedures of the studies available in Scopus, Web of Science (WoS), ProQuest, and EBSCO databases are presented in **Figure 1**. The entire process was conducted by two independent researchers, initially based on the title and abstract, followed by the procedure based on the full text of the study. In case of any disagreement between the assessing researchers, the third researcher was consulted to resolve the issue. Full texts of the studies were obtained from electronic databases or university libraries.

**Figure 1.**
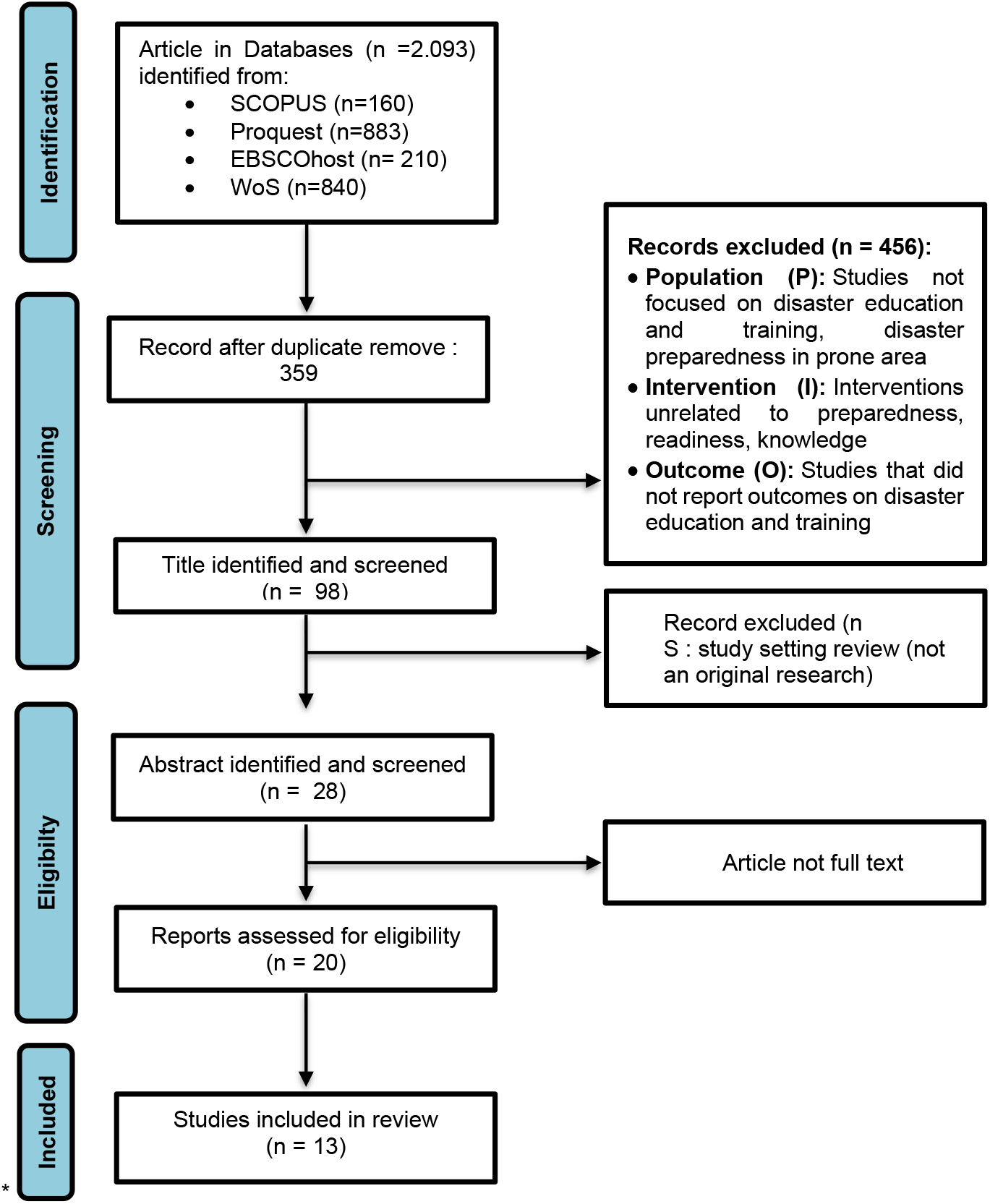
PRISMA Flow-chart. The identification, screening, and inclusion procedures of the studies were available in Scopus, Web of Science (WoS), ProQuest, and EBSCO databases.

A total of 2,093 records were identified from Scopus (n=160), ProQuest (n=883), EBSCO Host (n=210), and Web of Science (n=840). After duplicate removal, 359 articles remained. Following title and abstract screening, 98 were assessed, of which 28 underwent full-text evaluation. Finally, 13 studies met the inclusion criteria and were included in this review. The PRISMA flow diagram summarizing the screening process is presented in **Figure 1**. The 13 studies comprised a mix of randomized controlled trials (RCTs), quasi-experimental designs, cross-sectional analyses, pilot studies, and quality improvement projects, conducted in diverse settings across the Malaysia, Turkey, Nepal, Indonesia, China, and other regions. Sample sizes ranged from 62 to 3,675 respondents.

### Procedure of Data Extraction

Data extraction was conducted by two independent researchers. In case of any disagreement between the assessing researchers, the third researcher was consulted to resolve the issue. All necessary information was obtained from the full texts of the studies.

## RESULT

The results of the research articles on Disaster Education and Training Programs in Enhancing Community Preparedness Behaviour are presented in Table 1.

**Table 1.** Result of Article Analysis.

| No | Author(s),<br>Year | Objective | Design &<br>Sample | Intervention | Key Findings |
| --- | --- | --- | --- | --- | --- |
| 1 | (Pandya et al., 2024) | To evaluate the tabletop exercise approach in disaster preparedness and management at Nepal's first international conference | Longitudinal study, 140 Nepali healthcare professionals | Tabletop exercises on disaster leadership and communication | The tabletop exercise improved disaster preparedness and communication skills among healthcare professionals. |
| 2 | (Septikasari et al., 2024) | To analyze the urgency of disaster education in improving preparedness at elementary schools in disaster-prone areas | Quantitative, 264 elementary school teachers | Disaster education with three pillars: safe school facilities, disaster risk management, and disaster prevention education | 86% of schools had not implemented disaster education; Pillar 1 (safe facilities) was most urgent, followed by Pillars 2 and 3 (risk management and education). |
| 3 | (Yildiz et al., 2024) | To examine the effect of disaster education on children's risk perception and preparedness | Quasi-experimental longitudinal, 720 school children in Turkey | Disaster education on natural hazards and disaster risk reduction | Disaster education positively impacted children's risk perception, preparedness, and understanding of hazards. |
| 4 | (Lestari et al., 2025) | To evaluate the effectiveness of local wisdom-based disaster mitigation education in primary schools | Quasi-experimental, 346 students in Medan and Yogyakarta | Local wisdom integration in disaster education | Local wisdom integration significantly improved students' knowledge of disaster mitigation and preparedness. |
| 5 | (Noor et al., 2022) | To assess the impact of a Health Belief Model-based intervention on community flood preparedness | Cluster randomized controlled trial, community-wide implementation in six districts | Health Belief Model-based intervention (HEBI) | The intervention group showed significant improvements in flood preparedness, perceived benefits, and cues to action. |
| 6 | (Kalanlar et al., 2021) | To explore rehabilitation nurses' views on disaster | Quasi-experimental study, 200-bed hospital | Training and preparedness in disaster rehabilitation | Nurses reported moderate disaster preparedness, highlighting the |
|  |  | rehabilitation services and their preparedness |  |  | need for more training in disaster rehabilitation services. |
| 7 | (Adiyaman et al., 2025) | To evaluate the effect of earthquake preparedness training on mothers with physically disabled children | Randomized controlled trial, 78 mothers | Earthquake preparedness training | The training significantly improved mothers' psychological resilience and earthquake preparedness. |
| 8 | (Wang et al., 2023) | To investigate the mediating role of disaster education between school preparedness and student preparedness | Survey, 3,675 students in China | School preparedness and disaster education | Disaster education mediates the relationship between school preparedness and students' preparedness, enhancing their resilience. |
| 9 | (Erkin & Kiyan, 2025) | To assess the impact of integrating disaster nursing into nursing curricula on disaster literacy and preparedness | Quasi-experimental pretest-posttest study, 62 nursing students | Disaster Nursing course | Integration of disaster nursing improved students' disaster literacy and preparedness perceptions significantly. |
| 10 | (Mohammad et al., 2020) | To evaluate the effectiveness of community-based health education on preparedness for flood-related communicable diseases in Kelantan | Non-randomized community-controlled trial, 300 villagers in the intervention community (Tumpat) and 125 in the control community (Bachok) | Health education modules focusing on flood-related communicable diseases, including videos, health talks, and printed materials | Significant improvements in knowledge (9.4% to 52.6%), attitude (10% increment), and preventive practices, particularly in drinking safe water and protective habits. The intervention community showed higher knowledge compared to the control community. |
| 11 | (Batu et al., 2024) | To explore the most effective educational methods for conveying information | Cross-sectional and case-control, 405 students from Ege University, Turkey | Educational materials: brochures, face-to-face sessions, videos | Significant improvements in disaster knowledge after education, with video and face-to-face |
|  |  | about natural disasters in large institutions |  |  | methods proving more effective than brochures. Demographic factors like gender, faculty, and class year influenced knowledge levels. |
| 12 | (Sofyana et al., 2021) | To evaluate the effectiveness of the "Pokbaya Asalkena" model based on Transcultural Nursing in enhancing disaster preparedness in vulnerable communities | Mixed-method approach, 90 high school students from West Bandung, divided into three intervention groups | Training interventions using the Pokbaya Asalkena model with Transcultural Nursing approach | Significant increase in knowledge, attitudes, and skills related to disaster preparedness in all three intervention groups ($p = 0.000$). The Transcultural Nursing-based intervention showed the most improvement. |
| 13 | (Sofyana et al., 2024) | To implement and assess the effectiveness of the ILATGANA-PHN disaster preparedness training integration model to enhance community preparedness in disaster-prone areas | Pre-post test design, 78 community members from Kendeng Community, Sugih Mukti Village | ILATGANA-PHN training focused on disaster preparedness, emergency planning, and resource mobilization | The training significantly improved knowledge, attitudes, family emergency planning, and community resource mobilization, with $p\text{-values} \leq 0.005$ indicating strong effectiveness in increasing disaster preparedness. |

## DISCUSSION

This literature review comprises 13 articles that explore the effects of disaster education and training on disaster preparedness behaviours. Based on the analysis of 13 journals, the study used a quantitative method with a quasy experimental, cross-sectional stud, A pilot study methods which explores the influence of disaster education and training on community preparedness. Data collection was done through observation and experimental method. The sample in this study were individual students, nurses, and community who live in prone areas. The distribution of samples came from several countries including: Malaysia, Turkey, Nepal, Indonesia, China, and other regions. The effectiveness of disaster education and training in enhancing disaster preparedness has been a topic of growing interest in disaster management research. A cross various regions, disaster education models tailored to local contexts and needs have demonstrated substantial improvements in community preparedness. The studies examined in this literature review shed light on the diverse approaches to disaster training and their impact on disaster preparedness, with particular focus on the role of culturally sensitive methods and community empowerment.

Several studies underscore the significant role of tailored disaster education in enhancing individual, family, and community preparedness. For instance, Sofyana et al., (2021) investigated the effectiveness of the “Pokbaya Asalkena” model, a peer group empowerment model based on the transcultural nursing approach. This study found that targeted training, integrating local cultural contexts, significantly improved disaster preparedness among high school students in disaster-prone areas. The results indicated an increase in knowledge, attitudes, and disaster-related skills post-intervention (p = 0.000), highlighting the importance of culturally adapted educational programs. This aligns with previous findings by Sofyana et al., (2022), where culturally sensitive disaster risk reduction training was identified as a crucial factor for empowering communities in high-risk areas. The study emphasized that disaster preparedness should consider local culture and community dynamics, as culturally informed training programs are more likely to resonate with the population and foster active participation.

Similarly, the research by Yildiz et al., (2024) demonstrated that disaster education, particularly in school settings, plays a vital role in shaping children’s risk perceptions and preparedness. The study highlighted that children exposed to structured disaster education programs were better equipped to identify risks and take preventive actions. These findings are supported by Batu et al., (2024), whose study on disaster education methods suggested that video-based and face-to-face approaches were more effective than written materials. These interactive methods not only improved disaster knowledge but also motivated participants to engage actively in preparedness activities. The research by Sofyana et al., (2021) and Yildiz et al., (2024) also highlights the necessity of integrating disaster preparedness training into regular educational curricula, ensuring that students and community members are continuously exposed to the principles of disaster risk reduction.

A recurring theme in the literature is the positive impact of community-based disaster education, particularly through peer groups and local leadership. The work of Mohammad et al., (2020) and Sofyana et al., (2024) demonstrated that empowering peer groups, particularly through school children and youth, significantly enhances disaster preparedness. In both studies, peer educators were trained to serve as disaster preparedness leaders, increasing community engagement and creating a multiplier effect. This peer-driven approach fosters a sense of responsibility and preparedness within local communities, as participants are encouraged to share their knowledge and practices with others.

The ILATGANA-PHN model, developed by Sofyana et al., (2024), was particularly effective in enhancing community capacity by targeting local leaders and individuals in disaster-prone areas. The model showed substantial improvements in knowledge, attitudes, and skills related to disaster preparedness, highlighting the potential of community-based models in increasing disaster resilience. Moreover, the integration of disaster preparedness training with public health nursing, as seen in the ILATGANA-PHN model, reflects the growing recognition of the role of healthcare professionals in disaster risk reduction. Nurses, with their unique community access and health expertise, are well-positioned to lead disaster education initiatives and act as change agents.

Studies like those by Sofyana et al., (2024) reinforce the idea that nurses’ involvement in disaster preparedness training not only improves community preparedness but also strengthens the overall resilience of the healthcare system. The integration of training focused on emergency planning, resource mobilization, and disaster warnings was found to significantly enhance preparedness at both the individual and community levels (p = 0.000) (Mohammad et al., 2020; Lestari et al., 2025). One of the key insights emerging from these studies is the importance of continuous, structured training that extends beyond one-off interventions. A study by Batu et al., (2024) emphasized the need for ongoing engagement to maintain and reinforce disaster preparedness. Periodic training sessions, refreshers, and community drills help to keep disaster knowledge current and ensure that the community is ready to act in times of crisis. Moreover, disaster education’s role in improving psychological preparedness is crucial.

Research by Mohammad et al., (2020) and (Adiyaman et al., 2025) highlights the positive impact of training programs on participants’ psychological resilience. This study found that mothers of physically disabled children who participated in earthquake preparedness training showed significant improvements in their psychological resilience and preparedness levels. This suggests that disaster training not only increases knowledge and skills but also strengthens mental and emotional preparedness, which is essential for responding effectively to disasters.

In conclusion, disaster education and training are critical components of effective disaster risk reduction strategies. The studies reviewed highlight the positive impact of culturally sensitive, community-based training models in enhancing disaster preparedness. Moreover, the involvement of local leaders, schools, and healthcare professionals-particularly nurses-emerges as a key factor in increasing disaster resilience. While the evidence suggests that such interventions are effective, future research should focus on the long-term impact of disaster education on behaviour change, as well as explore strategies for overcoming barriers to engagement in disaster-prone areas. Further, more attention should be given to developing standardized, scalable models of disaster education that can be adapted to diverse cultural and geographical contexts.

### Implication

The findings reviewed emphasize the need for culturally sensitive and community-driven disaster preparedness programs. Culturally tailored models, such as the “Pokbaya Asalkena” approach, significantly enhance community engagement and effectiveness, suggesting that disaster education should be aligned with local cultural values. Furthermore, peer education and community empowerment have proven to be effective in spreading knowledge and fostering collective responsibility for disaster preparedness. This highlights the importance of integrating youth and local leaders into disaster education initiatives. The evidence also stresses the necessity for continuous disaster training. One-time interventions yield short-term improvements, but sustained engagement through periodic refreshers and community drills is crucial for maintaining long-term preparedness. Additionally, the involvement of public health nurses in disaster education is vital. Nurses, due to their trusted roles in communities, are well-positioned to lead preparedness initiatives, and policies should support their integration into disaster risk reduction efforts.

Finally, addressing the gaps in disaster education, particularly in rural and underserved areas, requires policy interventions that ensure equitable access to training. Standardized disaster preparedness programs must be developed and implemented, with active involvement from local authorities and community leaders to ensure their effectiveness. These findings underscore the need for disaster preparedness programs that are culturally relevant, community-based, and sustained over time to build stronger, more resilient communities.

### Limitation

Despite the valuable insights provided by the studies reviewed, there are several limitations that should be acknowledged in interpreting the findings and shaping future research in disaster preparedness education. First, many of the studies focused on relatively small sample sizes, which may limit the generalizability of the findings. For instance, the studies by (Sofyana et al., 2021, 2024) and (Batu et al., 2024) involved targeted interventions with limited numbers of participants, primarily from specific regions or institutions. While these studies provide valuable insights into disaster education methods, their findings may not be fully representative of broader populations, particularly in regions with different cultural or socioeconomic contexts.

Second, the majority of the studies relied on pre and post-intervention assessments to measure the impact of disaster education programs. While these designs are effective for evaluating immediate changes in knowledge, attitudes, and skills, they do not account for long-term behavioural changes or the sustainability of disaster preparedness beyond the intervention period. As noted in several studies, ongoing engagement and repeated exposure to disaster education are essential for maintaining high levels of preparedness, yet few studies have tracked the long-term effectiveness of these programs.

Finally, there is limited research on the scalability and adaptability of the disaster education models used in these studies. Most of the interventions were designed for specific local contexts or small-scale communities, and it remains unclear whether these models can be effectively scaled up or adapted to larger, more diverse populations. Further research is needed to test the transferability of these models to different geographic areas, cultural settings, and disaster contexts.

## CONCLUSION

The studies reviewed highlight the essential role of disaster education in enhancing community preparedness. Culturally tailored models, such as those based on Transcultural Nursing and peer-led empowerment, significantly improve disaster knowledge and engagement. These findings emphasize the importance of adapting disaster training to local contexts and involving communities in risk reduction efforts. Continuous education, rather than one-time interventions, is critical for maintaining long-term preparedness. However, gaps remain in understanding psychological barriers and the scalability of these models. Future research should focus on long-term impacts, psychological factors, and adapting disaster education to diverse settings. Overall, disaster preparedness education is vital for strengthening community resilience to natural disasters.

## Data Availability

All data produced in the present work are contained in the manuscript

